# Users’ Reactions on Announced Vaccines against COVID-19 Before Marketing in France: Analysis of Twitter posts

**DOI:** 10.1101/2022.02.14.22268832

**Authors:** Alexandre Dupuy-Zini, Bissan Audeh, Amandine Gagneux-Brunon, Cedric Bousquet

**Affiliations:** Sorbonne Université, Université Sorbonne Paris Nord, INSERM, Laboratoire d’Informatique Médicale et d’Ingénierie des connaissances en e-Santé, LIMICS, Paris, France; Centre International de Recherche en Infectiologie, Team GIMAP, Univ Lyon, Université Jean Monnet, Université Claude Bernard Lyon 1, Inserm, U1111, CNRS, UMR530, France; CIC INSERM 1408 Vaccinologie, CHU de Saint-Etienne, France; Chaire PREVACCI, Université Jean Monnet, Saint-Etienne, France; Service de santé publique et information médicale, CHU de Saint Etienne, France

**Keywords:** COVID-19, public health, Twitter, Vaccine, Social Media, Misinformation, France

## Abstract

**Background:** Within a few months, the COVID-19 pandemic has spread to many countries and has been a real challenge for health systems all around the world. This unprecedented crisis has led to a surge of online discussions about potential cures for the disease. Among them, vaccines have been at the heart of the debates, and have faced lack of confidence before marketing in France.

**Objective:** This study aims to identify and investigate the opinion of French Twitter users on the announced vaccines against COVID-19 through sentiment analysis.

**Methods:** This study was conducted in two phases. First, we filtered a collection of tweets related to COVID-19 from February to August 2020 with a set of keywords associated with vaccine mistrust using word embeddings. Second, we performed sentiment analysis using deep learning to identify the characteristics of vaccine mistrust. The model was trained on a hand labeled subset of 4,548 tweets.

**Results:** A set of 69 relevant keywords were identified as the semantic concept of the word “vaccin” (vaccine in French) and focus mainly on conspiracies, pharmaceutical companies, and alternative treatments. Those keywords enabled to extract nearly 350k tweets in French. The sentiment analysis model achieved a 0.75 accuracy. The model then predicted 16% of positive tweets, 41% of negative tweets and 43% of neutral tweets. This allowed to explore the semantic concepts of positive and negative tweets and to plot the trends of each sentiment. The main negative rhetoric identified from users’ tweets was that vaccines are perceived as having a political purpose, and that COVID-19 is a commercial argument for the pharmaceutical companies.

**Conclusions:** Twitter might be a useful tool to investigate the arguments of vaccine mistrust as it unveils a political criticism contrasting with the usual concerns on adverse drug reactions. As the opposition rhetoric is more consistent and more widely spread than the positive rhetoric, we believe that this research provides effective tools to help health authorities better characterize the risk of vaccine mistrust.

## Introduction

### Background

Since December 2019, the COVID-19 outbreak has led governments to impose a wide range of policies to help contain the effect of the pandemic. In France, after a 55-days total lockdown from March to May 2020, restrictions were partially lifted during the summer, but they were reinstated as soon as October 2020. Throughout the health crisis, the media reserved a high attention to treatments against COVID-19. For example, professor Didier Raoult, an internationally renowned French microbiologist [1], presented a study from the Institut Hospitalier Universitaire de Marseille, in which a treatment combining hydroxychloroquine and azythromycin made it possible to significantly decrease the viral load in patients with COVID-19 [2]. A part of the population was then doubtful of the reasons why health authorities did not authorize using these drugs. Believing that some therapeutic means are highly efficient to cure the disease makes it less desirable to apply preventive measures, which might have affected the perception of potential vaccines against COVID-19.

According to the WHO, “vaccine hesitancy refers to delay in acceptance or refusal of vaccines despite availability of vaccination services” [3]. As our study was performed before vaccines were available on the market in France, we privilege using the term “mistrust” rather than “hesitancy”, but we still use hesitancy when relevant. For example, we use the term “vaccine hesitancy” with the HPV vaccine as it was available on the market when findings about this vaccine were reported in the literature.

Questioning the relevance of vaccines has been a well-established trend in France since the end of the 19th century, when the hesitancy turned political. Vaccine hesitancy still focuses today on the opposition to compulsory vaccination, to the government intrusion into the practice of medicine, and the defense of individual liberties [4]. Arguments against vaccines are supported by different actors, and public sensitivity to these ideas has become a major political issue at both national and international levels. In France, a climate of skepticism about vaccines has been fueled by events such as the suspension of the hepatitis B vaccination on suspicion of side effects, and the issue of the H1N1 vaccination campaign where large expenditures were made but the epidemic turned out to be much less intense than expected. Another example that received major attention in the media worldwide is the association between vaccination and autism [5] supported by data [6] that was subsequently retracted. Before vaccines roll-out, 26% of the French population would refuse to be vaccinated if a vaccine against COVID-19 became available [7]. Previous attitudes of vaccine hesitancy were associated with negative attitudes toward COVID-19 vaccines [8]. Another study conducted by IPSOS October 8-13, 2020 [9] revealed that in France, 54% of respondents would get it if the vaccine was available. It was one of the worst scores among 15 countries with an average of 73%. IPSOS [9] conducted this study again at the end of 2020 from 17 to 20 December and showed that the “Total Agree” failed to 40%. Among 15 countries, France, was the most refractory to the vaccine against Covid-19. This was confirmed by a study of Lazarus et al. [10] that showed only 59% positive opinions about the vaccine in June 2020.

### Objective

In this paper, we explore the content of Twitter with the help of advanced machine learning techniques to identify the barriers and motivations concerning the announced vaccines against COVID-19 in France between January and August 2020. Our work aims to assess the characteristics of users’ opinions to identify positive and negative reactions about COVID-19 and reveal main elements related to vaccine mistrust in the COVID-19 context.

### Previous work on vaccines and social media

The period of the COVID-19 crisis favored the publication of numerous surveys on European citizens to assess vaccination intentions against COVID-19 and to identify the categories of individuals most susceptible to vaccine resistance [11]. These studies are based on controlled statistical methods where respondents are constrained by a limited number of answers pre-defined by the investigators [12]. As a result, reasons for vaccine hesitancy other than those proposed in the controlled studies cannot be detected. However, the public is exposed to new events on a daily basis, and additional reasons for vaccine mistrust may emerge rapidly and not be captured by static resources. Real-time monitoring of social media can be an indicator of society’s hottest emerging issues. As the exhaustive analysis of a large volume of messages is impossible, the use of recent advances in Natural Language Processing (NLP) becomes compulsory. Sentiment and opinion analysis regarding COVID-19 pandemic benefited from recent automatic approaches for social media analysis. For example, topic modeling was used on Twitter by Xueting et al. [13] to analyse public opinion towards COVID-19 in California and New York, and by Luo et al. [14] who performed a similar analysis on HPV vaccination. Hao et al. [15] used dynamic topic modeling to track governmental decision-making regarding risk, test, and treatment based on Tweets of U.S. governors and presidential cabinet members. Several infodemiology studies applied machine learning approaches to analyse social media. Daughton et al. [16] used supervised learning classifiers to identify human behaviors relevant to COVID-19. Similarly, Chen et al. [17] used dimension reduction and cluster analysis to support comparison between viral COVID-19 posts in Twitter and Sina Weibo, a non-English-speaking platform in China. In some cases, multiple artificial intelligence approaches can be used to construct an observation framework, such as [18] where a combination of several machine learning approaches is proposed, including natural language processing, word embeddings and Markov models to investigate COVID-19 related emotions.

Exploring vaccine hesitancy through online posts in social media is inspiring. While some studies focused on qualitative analysis of a limited number of posts [19] [20], others employed semi-automatic approaches such as [21] which used content and network analysis to study misinformation about HPV vaccine. Recently, the use of automatic approaches based on NLP has become frequent for quantitative study about vaccine hesitancy. For example, for sentiment analysis about HPV vaccine, [22] used topic modeling on discussion forums, and [23] used transfer learning on Twitter posts. Similar approaches were applied to analyse vaccine hesitancy in the COVID-19 context. [24] used deep learning and NLP to analyse public sentiment towards COVID-19 vaccine based on a set of Twitter and Facebook posts from the United Kingdom and the United States. [25] used topic models based on Latent Dirichlet Allocation [26] for sentiment analysis against COVID-19 vaccine among Australian twitter users. Such studies are specific to the cultural and political context that affected decision making in vaccination policy.

To our knowledge, most social media studies regarding COVID-19 vaccine hesitancy in France are qualitative, and do not benefit from advanced and efficient machine learning. Our approach relies on FastText [27], which can provide better performance when the vocabulary contains many syntactic or orthographic variations of the same word. We privileged transfer learning that allows to learn with a limited amount of data or low computational capacity.

There are now many pre-trained NLP models available, depending on the languages or texts used during training. Models in English were created to facilitate research, such as COVID-Twitter-BERT [28] which is trained on English tweets mentioning the COVID-19 pandemic. As such resources are not available in French, it is necessary to fine tune existing models for the classification of opinions on the announced vaccines before we may apply them to Twitter’s data.

## Methods

### Overview

This study was conducted in two steps that were designed to be accessible and easily adapted to any other subject at a specific time. The first step consisted in constructing a dataset of French tweets related to COVID-19. From this dataset, a subset of keywords relative to the word “vaccin” (vaccine in French) was selected. By restricting the dataset to the tweets containing at least one of these keywords, a dataset related to the topic of vaccine mistrust in French during the COVID-19 pandemic was reached. The second step focused on sentiment analysis. As this part requires machine learning, a small part of the restricted dataset needs to be hand labeled to fine-tune a pretrained model. After training and evaluating the model, label prediction was performed on the whole restricted dataset and the predictions were explored in terms of vocabulary and timeline. These steps are summarized in Figure 1 and explained in detail in the subsections hereafter.

**Figure 1.**
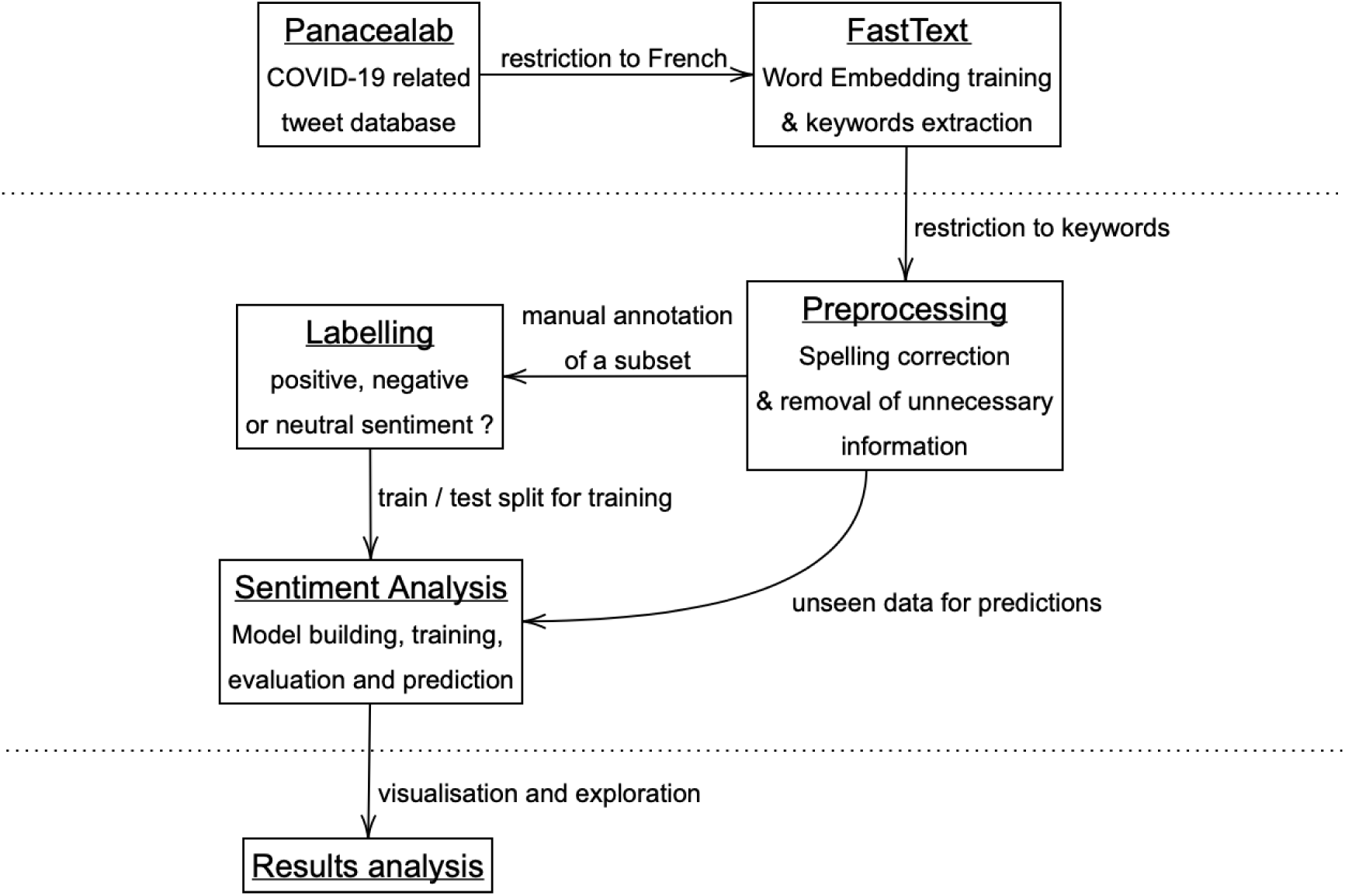
Summary of the study methodology, with the different phases separated by horizontal lines.

### Data Collection

Data were collected before vaccines were available on the market. Between February and August 2020, about 700 million unique tweets were identified by the PanaceaLab team at Georgia State University [29] using a selection of keywords that are mainly COVID-19 designation variants, and the corresponding Ids were made available on GitHub. A new version is made available every week and collects approximately 3 million tweets per day. Once restricted to tweets in French, the database has yet to be restricted to the vaccination topic.

On this purpose, word embeddings were trained on the whole French tweet dataset. Word embeddings are a way of representing a word of the vocabulary in a mathematical space. Words are transformed into vectors of a fixed number of dimensions to embed information about their meaning in the corpus. Thus, words that are close in meaning will be close in distance in this space. The choice of the word embeddings model is based on the properties and specificities of the data, as it influences the performance of the algorithms using them. As this pandemic period is exceptional, some words have emerged, and others have quickly changed meaning. The use of pretrained word embeddings does not make sense since they won’t highlight the link between vaccines and hydroxychloroquine in our specific context for example. Indeed, word embeddings were trained before the first case of COVID-19 was reported and vaccines against COVID-19 were not available at this time. Additionally, the corpora used for training these embeddings did not mention that hydroxychloroquine was being considered as a treatment for COVID. For this reason, we trained our own word embeddings model in order to be specific to the content present in the tweets at the time of the study, and take into account associations between concepts relative to the pandemics. Since the data is composed of short, noisy messages with uncertain spelling, the word embeddings generated by FastText offer significant advantages. Notably, this method is considered to be fast and enables words with a similar spelling to be brought together by using parameter sharing. Most word embedding models learn a vectorial representation from the word’s context, but as FastText also learns additional embeddings at the character level of the word, the decomposition of an unknown word enables to learn more relevant embeddings for rare words. The aforementioned technique named parameter sharing is an advantage since most models do not address the diversity of morphologically rich languages, such as French. Accurate word representations are difficult to learn since many word forms occur too rarely in the training corpus. Parameter sharing also enables to handle the uncertain spelling observed in tweets.

As word embedding models come with hyperparameters, tuning is necessary to fit the corpus at best. The metric used for that task is a criterion named the Discounted Cumulative Gain (DCG), which considers a user-defined collection of word pairs that are known to be close in meaning or context and computes the score of the cumulative closeness of the embeddings associated with the pairs. The higher this metric is, the more the model is expected to fit the corpus.

To complete the restriction of the French tweets to a vaccine centered dataset, a semantic field of the word “vaccin” is built: by iterating from the word “vaccin”, words close in the embedding space were added to the semantic field by computing their distance. The final restricted dataset is then composed of tweets containing at least one of those words, and the keywords identified were also used to explore the topics surrounding the vaccination topic.

### Data Preprocessing

Data preprocessing was designed to be as minimal as possible and focused on two main tasks. The first one was to delete unnecessary information and the second was to lower the noise in texts.

Firstly, the following steps were applied to the restricted dataset: URLs, punctuation, and special characters were removed. Tweets were then split into lists of lowercased words, and words that do not add any real value to the meaning (Stop Words) are also removed. In addition, some words such as “hydroxychloroquine” have many spelling variations. In order to suppress a part of the noise to obtain better performances, a spelling correction step was applied. We observed that correcting the 3 000 most common mistakes could correct around 70% of the misspelled words in the corpus. Thus, a dictionary of 3,482 words was manually created to correct this noise.

Finally, one twitter user appeared to be likely a bot by repeatedly posting 3 identical texts mentioning different users each time. It represented a total of 2,343 tweets, published in a very short period. This user was therefore deleted in order not to bias the results of the study.

### Model Building and Training

After the preprocessing steps, an extended analysis to explore users’ sentiments about vaccination was initiated using advanced machine learning models. The current state of the art in NLP does not allow for an unsupervised method classifying sentiment around the topic of vaccination. Nevertheless, it is fairly straightforward to access pre-trained models in French that can be adapted to a supervised classification task. Thus, as the model already has a sufficient understanding of the French language, it is not necessary to attain a large quantity of hand labelled tweets.

Most recent language models are based on the BERT model [30]. Several pre-trained models are available, with different sizes of architecture, which are specific to one or more languages, or which are trained on a particular type of text. To use these models for sentiment analysis, we chose to construct a classifier at the output of a BERT-type model to determine which one of the classes is the most likely for each tweet. The BERT-type model chosen here is CamemBERT [31], to which a linear classification layer is added. This linear layer learns the best multidimensional linear regression to perform on the output of the BERT-type model to obtain the desired predictions. Many other classification layers can be evaluated to improve performance. Moreover, this implies that the training is supervised.

This study classifies tweets into 3 categories: positive, negative, or neutral sentiment. Tweets labeled as positive mention the announced vaccine in an optimistic, confident way and often diffuse encouraging news on the subject. The negative ones evoke the potential vaccine in a mistrustful, and possibly conspiratorial way, and they seek to warn of its possible risks or manipulation or to relay information spreading doubt about its effectiveness. The neutral ones are unrelated to vaccines or do not contain any judgment about them.

For training, a total of 4,548 tweets (1.3% of the restricted dataset) were labelled, of which 26.9% were negative, 21.2% positive and 51,9% neutral. The classification task was therefore imbalanced, and this had to be addressed by using specific methods. Firstly, a stratified training method was required so that the training and test sets respect the proportion of each class. In order to compare methods, metrics must ensure that each class is best predicted not only as a whole, but also class by class. Precision (ratio of predicted items that truly belong to this class), and recall (ratio of correctly predicted items among the known items of this class) were used to measure the performance of our method class by class, in addition to the F1-score, which is their harmonic mean. Accuracy (ratio of correct predictions) is the metric used to give a general appreciation of the performance of the models.

### Ethics

The Ethics Committee of the “CHU de Saint-Etienne” has given a favorable opinion on the conduct of this study, and referenced the project under the number IRBN1412020/CHUSTE. This opinion was motivated by the fact that the study is based only on data extracted from Twitter which are openly accessible to the public.

## Data & Materials availability

The original dataset providing the tweets’ ids is to be found on the Panacealab website https://github.com/thepanacealab/covid19_twitter.

## Results

### Collected Data

The initial dataset restricted to French tweets consisted of 894,315 unique words for 4,020,525 tweets. After training word embeddings of 300 dimensions, relations between words were explored, and 69 keywords were identified as the semantic concept of the word “vaccin”. Using Principal Component Analysis (PCA), we displayed a projection of the vector representations of each keyword on a shared two-dimensional plan (Figure 2). This projection seems to group keywords into 3 clusters surrounding the word “vaccin”, which could be summarized as: potential treatments (top left), conspiracy (bottom left), and pharmaceutical companies (right). The projection of the terms “vaccination” and “vacciner” (to vaccinate, in French) sets these words among the ones related to conspiracy theories terms like “complot” (conspiracy), “puce” (chip), and mentions of Bill “Gates” and George “Soros”. Although “vaccination” and “vacciner” have the same root as “vaccin”, the embeddings captured nuances in their contexts. This information already unveils some polarity that is associated with the vocabulary.

**Figure 2.**
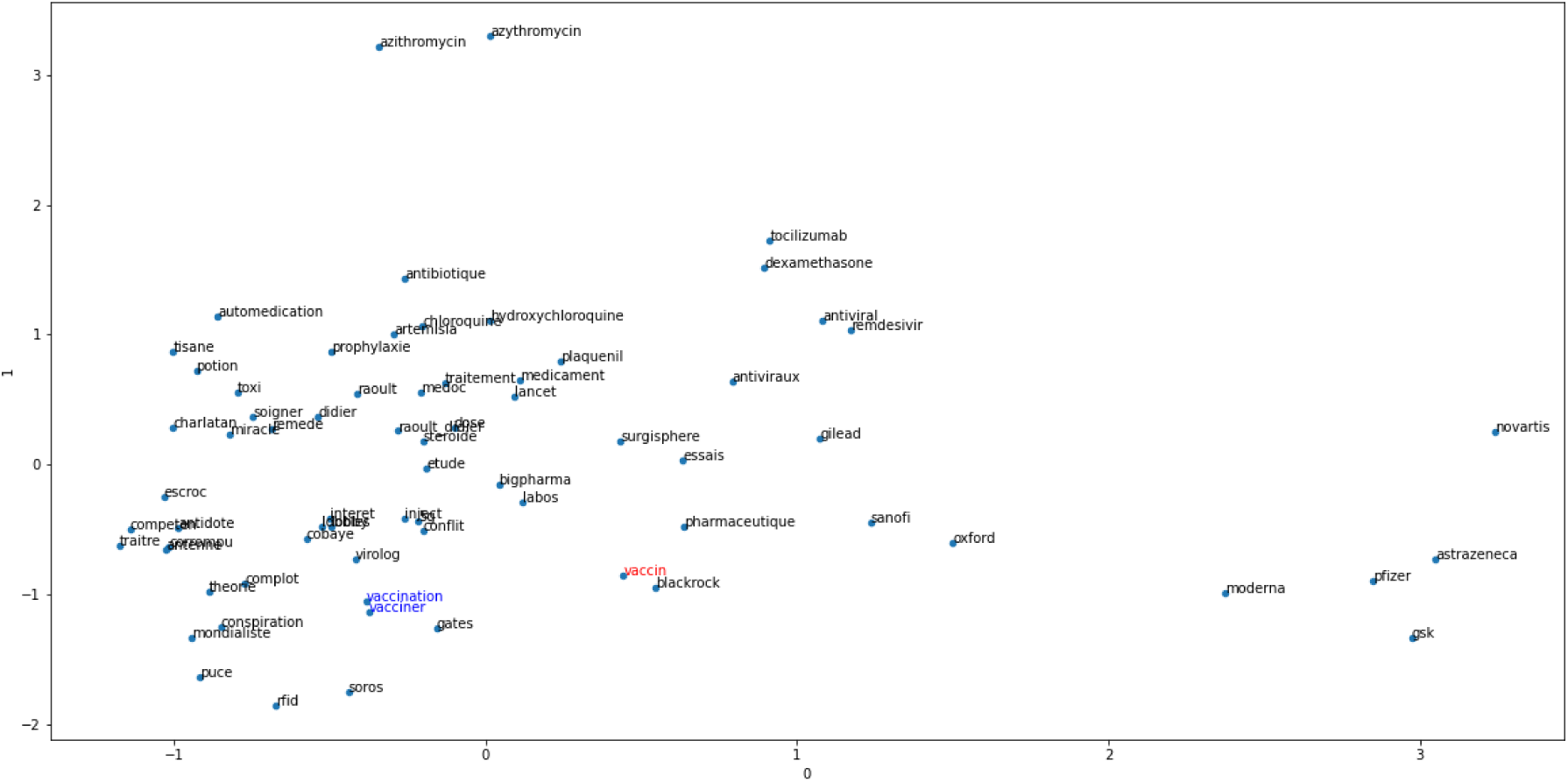
Two-dimensional projection of the 69 identified keywords, with the starting word “vaccin” in red, and “vaccination” and “vacciner” in blue for readability reason.

Tweets filtering based on word embeddings provided a specific dataset by restricting the collection to 344k tweets in French (around 9% of the initial French dataset). Filtering based on the “vaccin” word alone would have generated a subset of 75k tweets, thus ignoring a large number of tweets potentially related to aspects of vaccine mistrust. The results presented in the rest of this paper concern the 344k French tweets related to a potential vaccine against COVID-19.

### Model Evaluation

To be able to compare the performance of a more complex model using CamemBERT and a linear layer classifier, it is necessary to evaluate very simple models and to know their performances. Two simple baseline models were chosen. Since the classes are very imbalanced in favor of the neutral class, the simplest model is to predict only this neutral class. This first baseline model has an accuracy of 0.52, and any other model must get better performances than this. The second baseline model is more meaningful while still being very simple. It combines a document embedding vectorization to tf-idf features and a classifier named Multinomial Naïve Bayes. A document embedding is a vector representation of each document, according to the words of which it is composed. It is then used by the classifier to predict one of the three classes. After training, the metrics of this baseline are summarized in the following table (Table 1). While the precision of the model in predicting a positive or a negative tweet is 0.98 and 0.91 respectively, the recall falls to 0.19 for the positive tweets and 0.56 for the negative ones. Moreover, the prediction of the neutral class has a high recall of 0.99 but a low precision of 0.64 compared to the precision of positive and negative tweets prediction. Positive and negative tweets seem to be misclassified into the neutral class, that has therefore a weaker precision. The confusion matrix confirms those conclusions (Figure 3).

**Table 1.** Classification summary for tf-idf and Multinomial Naïve Bayes (MNB), and CamemBERT with the linear layer

|  | precision |  | recall |  | F1-score |  |
| --- | --- | --- | --- | --- | --- | --- |
|  | MNB | CamemBERT | MNB | CamemBERT | MNB | CamemBERT |
| positive | 0.98 | 0.64 | 0.19 | 0.73 | 0.32 | 0.68 |
| negative | 0.91 | 0.75 | 0.56 | 0.77 | 0.69 | 0.76 |
| neutral | 0.64 | 0.83 | 0.99 | 0.75 | 0.77 | 0.78 |
| macro average | 0.84 | 0.74 | 0.58 | 0.75 | 0.59 | 0.74 |
| micro average | 0.78 | 0.76 | 0.70 | 0.75 | 0.65 | 0.75 |

**Figure 3.**
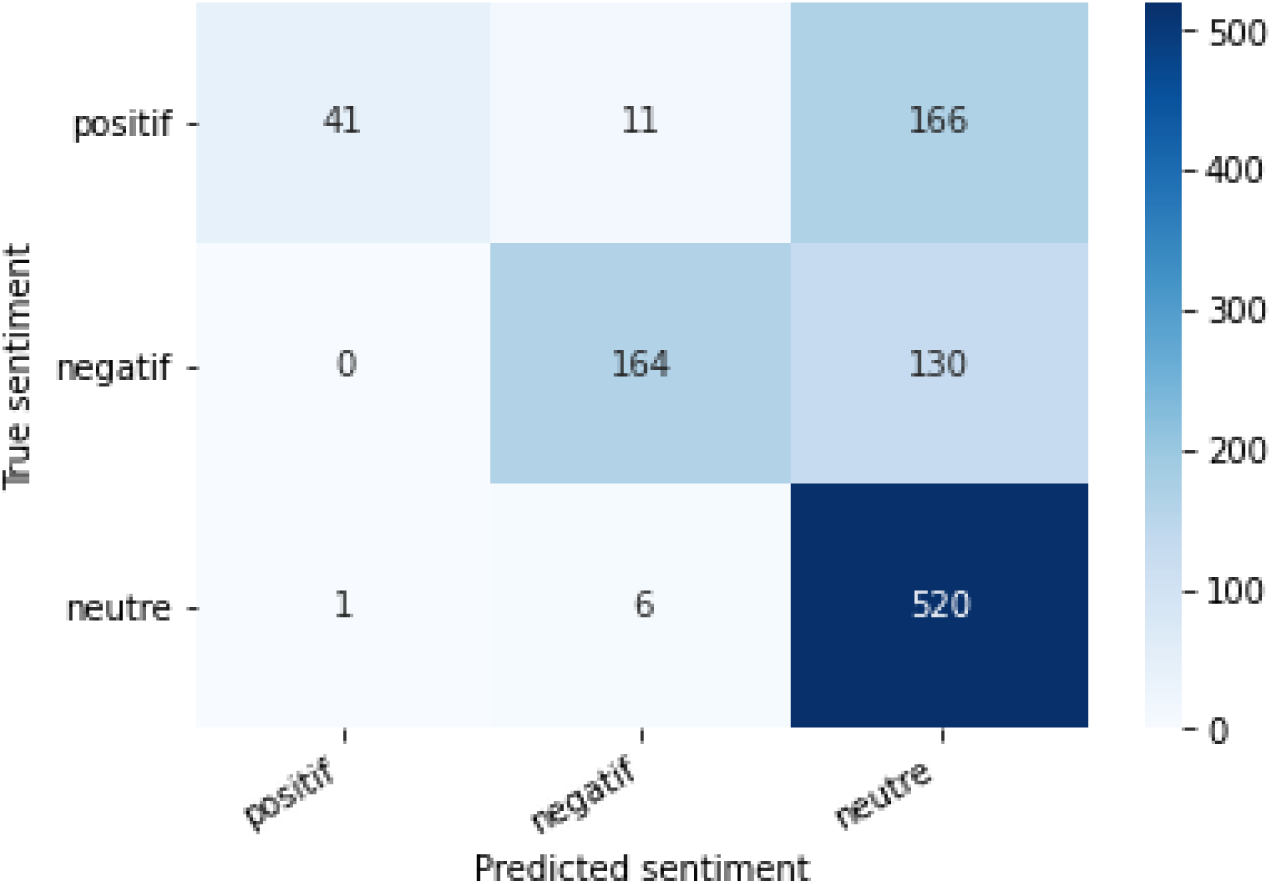
Confusion matrix for tf-idf and Multinomial Naïve Bayes

The performance of the sentiment analysis model proposed in this paper, which combines CamemBERT and a linear layer, is also summarized in table 1. All F1-scores are higher than for the baseline. The proposed model therefore obtained less misclassification for each class than the baseline. This conclusion is particularly visible for positive tweets predictions, of which there are less missed true positive (recall). Those results are also confirmed by the confusion matrix (Figure 4). However, the margin of error is still rather high as the accuracy is only 0.75 compared to 0.70 for the Multinomial Naïve Bayes model, but it is important to keep in mind the ambiguity of most tweets, whose annotation is subject to personal interpretation.

**Figure 4.**
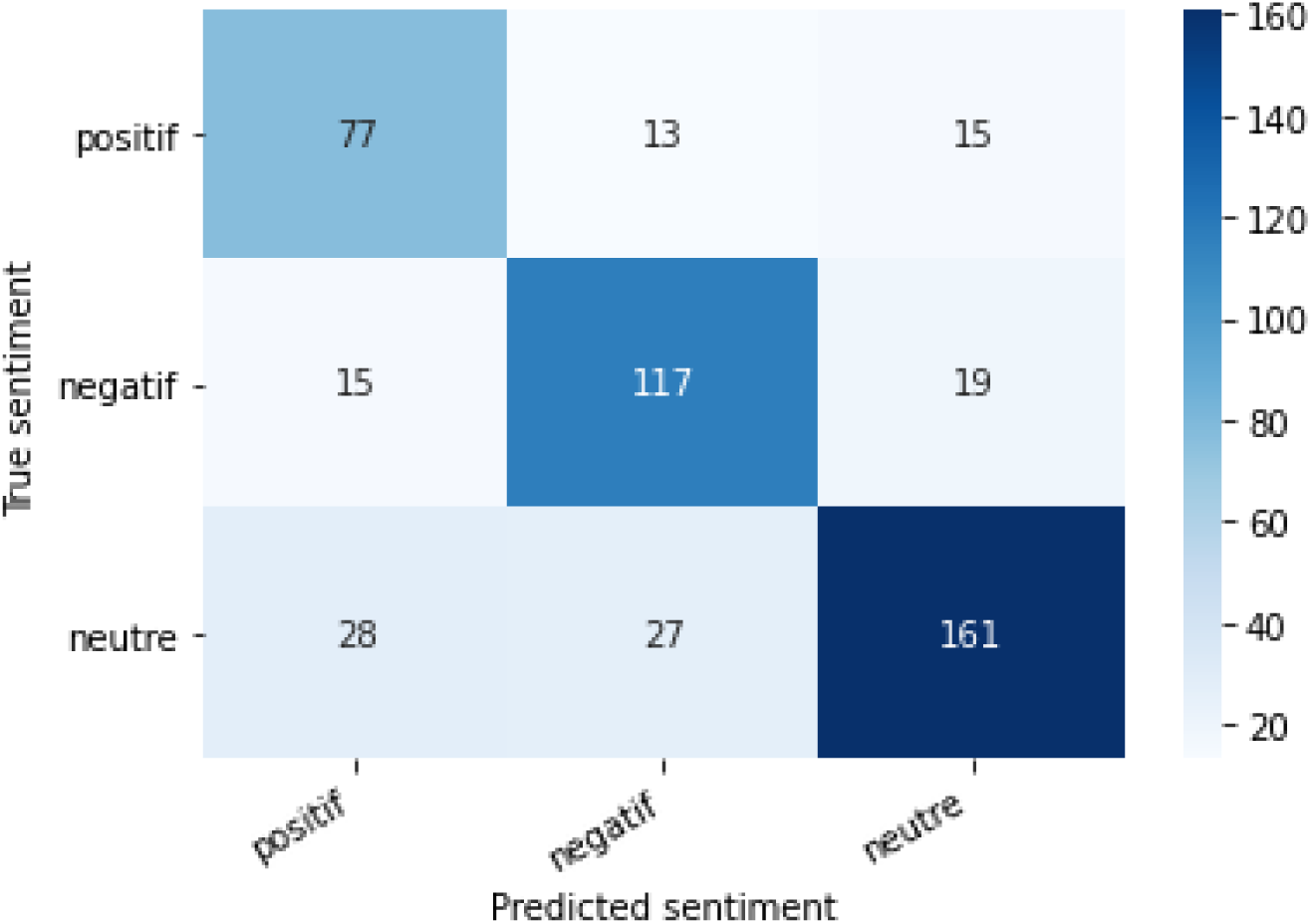
Confusion matrix for CamemBERT and a linear layer.

### Characteristics of Vaccine mistrust

In order to summarize the information obtained from this classification, two representations are chosen: the temporal presentation of the tweet counts per class, and the presentation of the most common words for each sentiment. As neutral tweets provide little information on the reasons of vaccine mistrust, the visualizations will only focus on the positive and negative tweets. Figure 5 displays the count of positive and negative tweets per day during the period of the study. Word embeddings using FastText allowed to identify a broader range of arguments for vaccine mistrust. However, this method included more negative opinion than positive opinion, compared to the counts per day if only tweets containing the word “vaccin” were considered. The wider the vocabulary is, the weaker signals of vaccine mistrust are detected.

**Figure 5.**
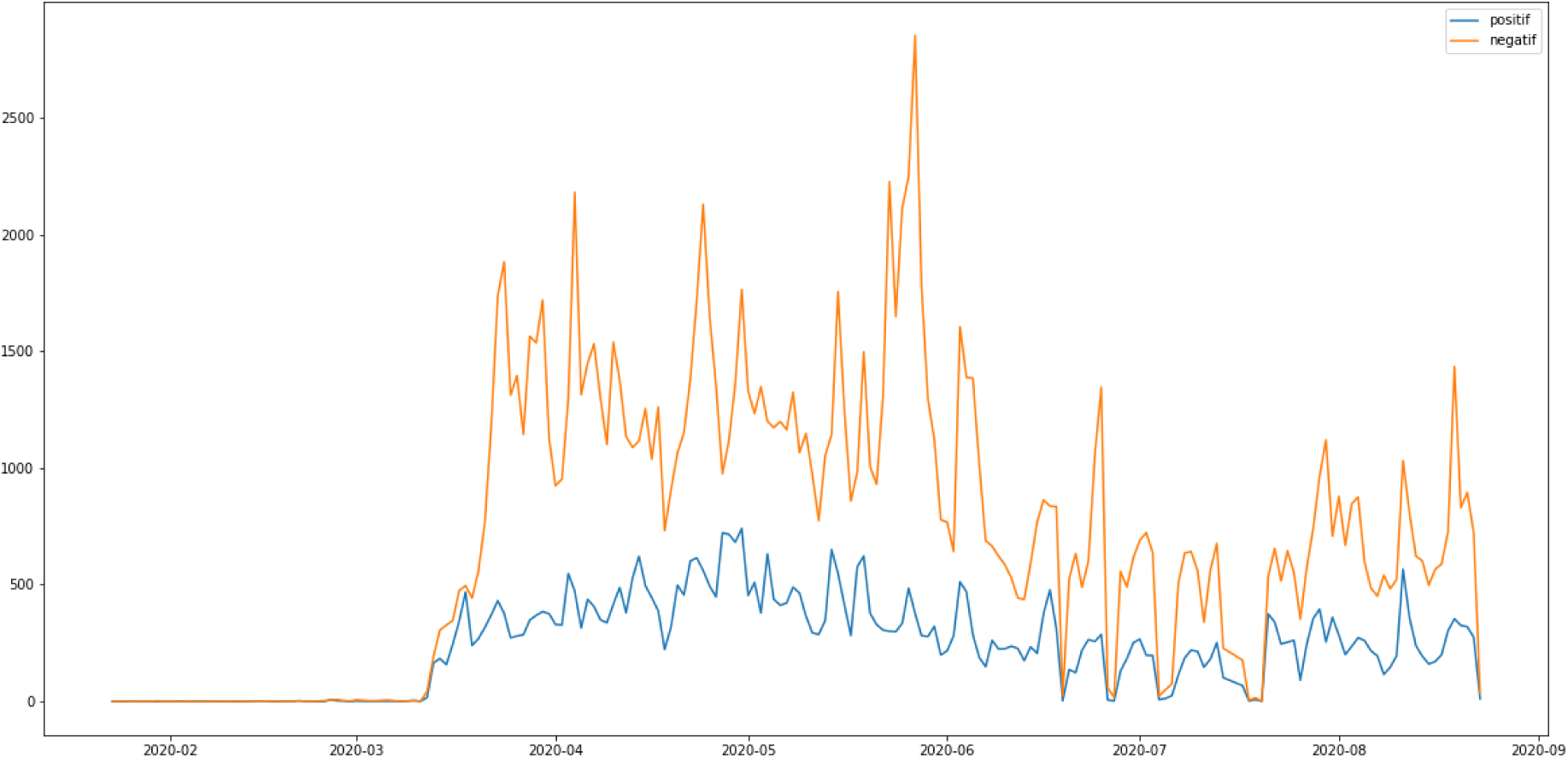
Counts of positive (blue) and negative (orange) tweets per day during the period of the study.

It is thus possible to distinguish days with a higher activity than usual for both sentiments. We assume that evaluation should focus on days when counts of tweets showing positive or negative sentiment show a polarization of the users’ opinions, rather than days where the numbers of tweets is high but mostly neutral. As trends sometimes fluctuate a lot from one day to the next, there seem to be temporary events which users quickly take advantage of.

Simultaneously, it is possible to explore the most used vocabulary per sentiment. The following figures present the words (Figure 6) and bigrams (Figure 7) that are most frequent per sentiment. Discourses are more homogenous in the negative tweets, where they focus on alternative treatments (“hydroxychloroquine” …), political contestation of the government (“désobéissance civile”, “gilets jaunes” …) (civil disobedience, yellow vests named after the yellow high-visibility vests worn by protesters during a movement that emerged in France in October 2018) and conspiracies (“bill gates”, “boycott cac40” …). This homogeneity is an advantage to isolate these negative discourses. Meanwhile, discussions about research advances are among the most common in the positive tweets. The most frequent positive terms focus on the efficiency or early results of the candidate vaccines (“efficace contre”, “résultats encourageants”…) (“Effective against”, “Encouraging results”). Nevertheless, positive tweets also communicate on some terms like “chloroquine” which are mostly attributed to negative tweets.

**Figure 6.**
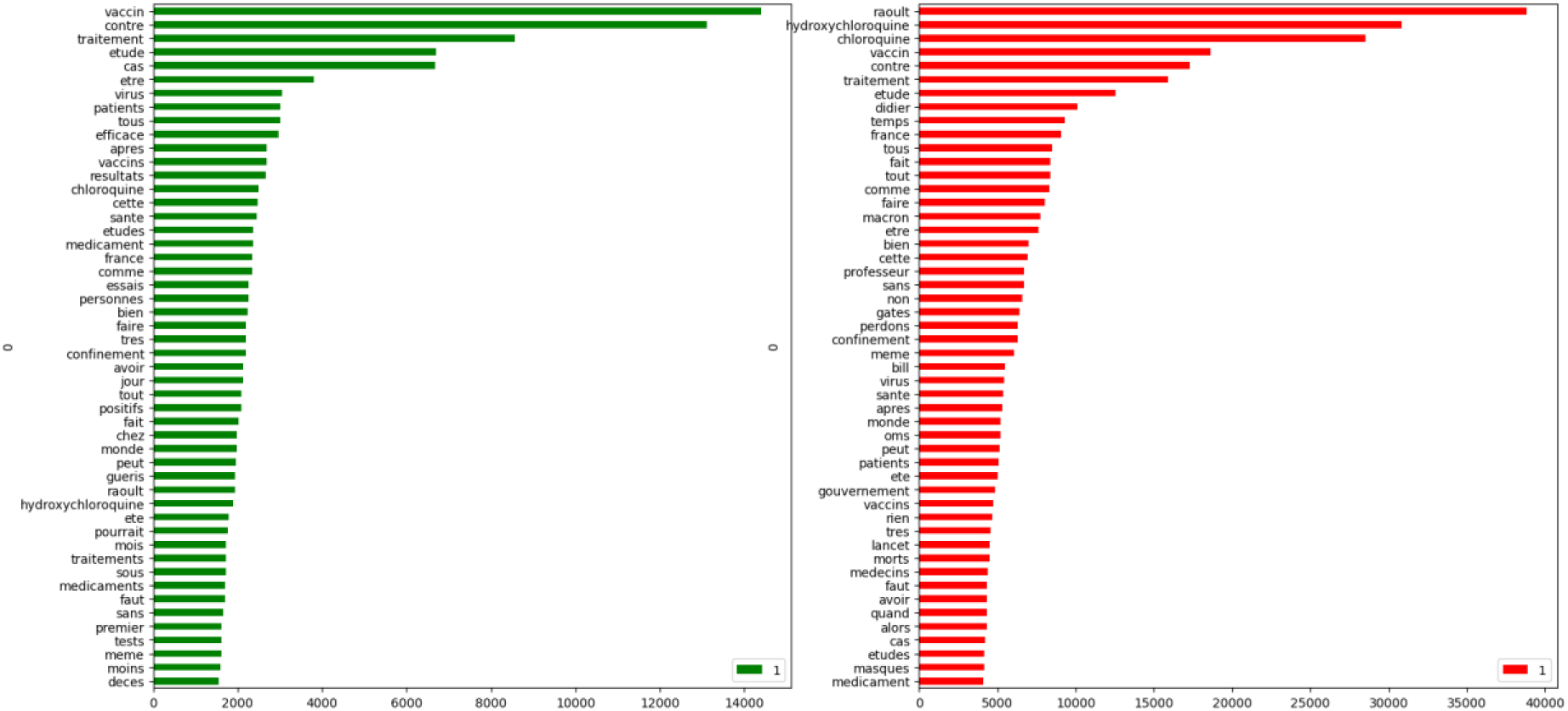
Most frequent word per sentiment, where green is positive and red negative.

**Figure 7.**
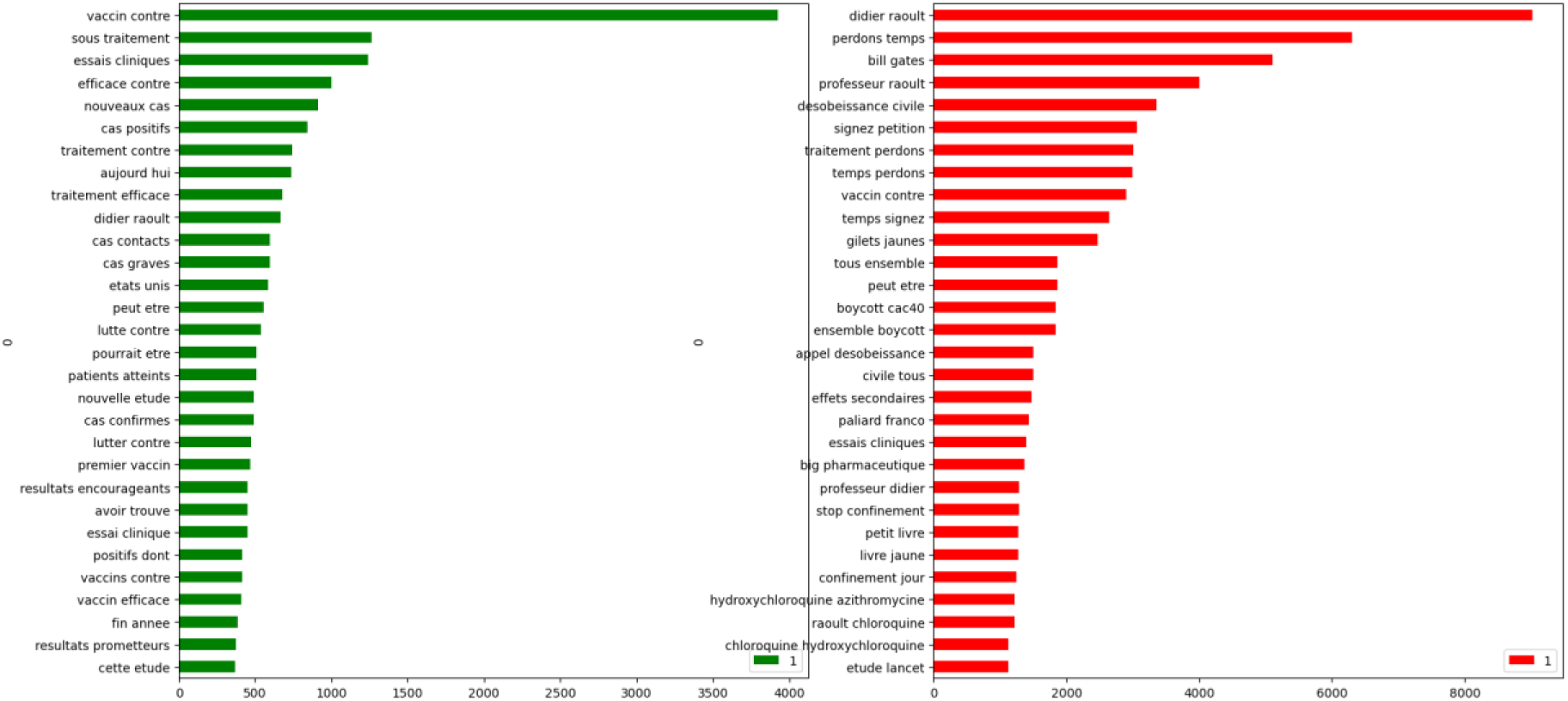
Most frequent bigrams per sentiment, where green is positive and red negative.

## Discussion

### Main Results

The objective of this study was to identify the opinion of Twitter users on the subject of a potential vaccine against COVID-19. After a review of the literature in both the medical and machine learning domains, the steps chosen were first to extract a set of relevant tweets, then to exploit them from a sentiment analysis perspective, and finally to explore these results. Using a vectorial vocabulary representation method proved to be a powerful way of broadening the lexical concept of vaccination during this pandemic. The PCA in figure 2 showed that “hydroxycloroquine” is close to “treatment” and “Raoult”, the doctor who was the first to promote this drug for the management of COVID-19. “Vaccination” is a neighbor of these terms which confirms our assumption that perception of vaccines should be considered in relation of perception of hydroxychloroquine. However, it does not allow to evaluate if believing a treatment is efficient to heal quickly may influence the intention of individuals to protect themselves because the proximity relations on the PCA are not typed. Nevertheless, further experiments in more recent periods could help to figure if such relations should also be considered between vaccines and other presumed treatment such as ivermectin [32] or azithromycin [33]. Our analysis revealed the dominant discourses and weak signals of vaccine mistrust.

After annotating a subset of tweets, an efficient classification implementing the state of the art of NLP was able to reveal temporal trends in sentiment about a potential vaccination against COVID-19. Moreover, further explorations of the vocabulary gave a different view of vaccine mistrust arguments.

Hence, there appears to be a change in the profile of Twitter users on this issue. According to Massey [34], Twitter seemed to be used more by profiles confident in the Human Papillomavirus Vaccine, whereas our study showed a greater sharing of opposition against the announced COVID-19 vaccine, notably driven by political distrust. Skepticism already observed with other vaccines could influence people that remain uncertain to be vaccinated because of the large audience that distrustful tweets have. Those observations may help the regulatory authorities to disseminate credible information by providing a clear and precise communication around a potential campaign. The success of a vaccination campaign depends not only on the sufficient coverage of the population to obtain collective immunity, but above all on the acceptance of such a campaign by the same population. This study tends to clarify reasons of vaccine mistrust based on users’ reactions on social media.

Finally, it is important to emphasize that this study is not representative of either the French population or Twitter users in general.

### Limitations

This study focuses on exploiting the textual information of Twitter but doesn’t extract any further metadata such as users’ information. However, a preliminary experience that we conducted earlier showed that the medical professionals seem to be excluded of the debate on social networks, except for a few personalities who are against a potential vaccine. This could lead to a better understanding of the observed dynamics.

Another limitation relies on the performances of the sentiment analysis. The model could achieve better performances in the near future with better parameters’ optimization and further exploration of other approaches. Models that are unsupervised like zero-shot learning could be interesting for additional investigations.

### Related work

Mistrust about COVID-19 vaccine has spread widely across social media. Consequently, its influence was able to reach a large part of the population. This mistrust situation was causing concern for health authorities, including the WHO, which lists vaccine mistrust as one of the 10 biggest threats on global health in 2019 next to the threat of a pandemic [35].

According to [36], there are many reasons for this mistrust: One may be doubtful of the vaccine benefit, there may be concerns about long term unexpected side effects, marketing of vaccines may be considered as a mere commercial operation where vendors are profiteering from patients, and one may have a preference for natural immunity rather than getting immunity from the vaccine. Other studies have considered conspiracy theories as an element influencing the decision of getting vaccinated [37]. Examples of these theories in the context of COVID-19 are Bill Gates’ intention to create a “global surveillance state” [38], and the economic motivation of the “Big Pharma” vaccine industry [39].

Analyzing social media can facilitate the evaluation of the adherence to a potential COVID-19 vaccine. Our analysis of Twitter for the period from February 1^st^ to August 25, 2020 has shown that a large share of negative posts mention chloroquine / hydroxychloroqine and Professor Didier Raoult. As we mentioned in the background, a part of the population has doubts about the non-authorisation of this treatment. These doubts could have been supported by the highly mediatised “Lancet Gate” in which the WHO urgently stopped trials on using chloroquine and hydroxychloroquine as a treatment against COVID-19 based on scientific papers that used corrupted data and that were retracted shortly later [40] [41].

The high accessibility to social media today brings to the conclusion that popular news on this resource can reach an important number of people in a little time [42]. The general director of the World Health Organization declared in 2020 that the WHO must deal with the infodemic in addition to the pandemic [43]. A new report published by the Centre for Countering Digital Hate (CCDH) noted that 31 million people follow anti-vaccine groups on Facebook, with 17 million people subscribing to similar accounts on YouTube [44]. Such accessibility to a large volume of information can help identify public views. However, this resource cannot replace more controlled survey methods because of the inherent selection bias on one hand, and the uncontrolled spread of false information in this resource on the other hand [45].

## Conclusions

Our study showed that Twitter could be a useful tool to investigate the arguments of vaccine mistrust. Our results unveil that political aspects of vaccination overshadow its usual criticisms about adverse drug reactions. As the opposition rhetoric is generally more homogenous and more widely spread than the positive rhetoric, we believe that this study provides effective tools to help health authorities better understand vaccine mistrust.

## Data Availability

The original dataset providing the tweets ids is to be found on the Panacealab website.

https://github.com/thepanacealab/covid19_twitter

## Acknowledgements

ADZ implemented the embeddings and the classifier, and performed the analysis. BA collected data from the Panacelab github repository and extracted tweets from github, administered the server, and installed the necessary libraries and tools. ADZ and CB drafted the manuscript. AGB provided support on designing the study and reviewing results. All co-authors revised the article critically for important intellectual content and provided final approval of the version to be submitted.

This work was funded by the grant N° ANR-16-CE23-0011-01 from the ANR, the French Agence nationale de la Recherche through the PEGASE (Pharmacovigilance enrichie par des Groupements Améliorant la detection des Signaux Emergents) project. Exaion, a new subsidiary of the EDF group, a cloud provider of blockchain and high-performance computing solutions, lent one of its servers with two graphical processing units free of charge for the duration of the study. We thank the representatives of Exaion who helped us: Fatih Balyeli, Laurent Bernou-Mazars, Nicolas Meaux, and Vivien Sayve. We acknowledge the work of the Panacea Lab in collecting tweets related to the COVID-19 pandemics. The views mentioned in this article are those of the authors only.

## Abbreviations

BERT: Bidirectional Encoder Representations from Transformers
COVID-19: coronavirus disease
NLP: Natural Language Processing
Tf-idf: Term Frequency – Inverse Document Frequency
WHO: World Health Organisation

## References

[1] D. Raoult, “CORONAVIRUS : VERS UNE SORTIE DE CRISE ?,” 25 February 2020. [Online]. Available: https://www.mediterranee-infection.com/coronavirus-vers-une-sortie-de-crise/. [Accessed August 2021].

[2] P. Gautret, J.-C. Lagier, P. Parola, V. T. Hoang, L. Meddeb, M. Mailhe, B. Doudier, J. Courjon, V. Giordanengo, V. E. Vieira, H. T. Dupont, S. Honoré, P. Colson and Cha, “Hydroxychloroquine and azithromycin as a treatment of COVID-19: results of an open-label non-randomized clinical trial,” International Journal of Antimicrobial Agents, 2020.

[3] N. E. MacDonald, “Vaccine hesitancy: Definition, scope and determinants.,” Vaccine, vol. 33, pp. 4161–4164. doi: 10.1016, August 2015.

[4] J. Ward, C. Alleaume, P. Peretti-Watel, V. Seror, S. Cortaredona, O. Launay, J. Raude, P. Verger, F. Beck, S. Legleye and O. L’Haridon, “The French public’s attitudes to a future COVID-19 vaccine: The politicization of a public health issue,” Social science & medicine, vol. 265, p. 113414, 2020.

[5] A. Sabra,. A. B. Joseph and. R. C. Angel, “Ileal-lymphoid-nodular hyperplasia, non-specific colitis, and pervasive developmental disorder in children,” The Lancet, pp. 234–235, 1998.

[6] B. Deer, “How the case against the MMR vaccine was fixed.,” Bmj, 2011.

[7] P. Peretti-Watel, V. Seror, S. Cortaredona, O. Launay, J. Raude, P. Verger and J. K. Ward, “A future vaccination campaign against COVID-19 at risk of vaccine hesitancy and politicisation.,” The Lancet Infectious Diseases, pp. 769–770., 2020.

[8] M. Detoc, S. Bruel, P. Frappe, B. Tardy, E. Botelho-Nevers and A. Gagneux-Brunon, “Intention to participate in a COVID-19 vaccine clinical trial and to get vaccinated against COVID-19 in France during the pandemic,” Vaccine, vol. 38, no. 45, pp. 7002–7006, 2020.

[9] Ipsos, “Global attitudes on a COVID-19 vaccine,” 2020. [Online]. Available: https://www.ipsos.com/sites/default/files/ct/news/documents/2020-11/global-attitudes-on-a-covid-19-vaccine-oct-2020.pdf. [Accessed 7 2021].

[10] J. V. Lazarus, S. C. Ratzan, A. Palayew,. O. G. Lawrence, H. J. Larson, K. Rabin, S. Kimball and A. El-Mohandes, “A global survey of potential acceptance of a COVID-19 vaccine,” Nature Medicine, 2021.

[11] S. Neumann-BÖhme, N. E. Varghese, I. Sabat and et al., “Once we have it, will we use it? A European survey on willingness to be vaccinated against COVID-19.,” The European Journal of Health Economics, vol. 21, p. 977–982, 2020.

[12] D. Freeman, B. S. Loe, L.-M. Yu, J. Freeman, A. Chadwick and C. Vaccari, “Effects of different types of written vaccination information on COVID-19 vaccine hesitancy in the UK (OCEANS-III): a single-blind, parallel-group, randomised controlled trial,” The Lancet Public Health, vol. 6, no. 6, pp. 416–427., 2021.

[13] W. Xueting, Z. Canruo, X. Zidian and L. Dongmei, “Public Opinions towards COVID-19 in California and New York on Twitter,” 2020. [Online]. Available: https://www.medrxiv.org/content/10.1101/2020.07.12.20151936v1.

[14] X. Luo, G. Zimet and S. Shah, “A natural language processing framework to analyse the opinions on HPV vaccination reflected in twitter over 10 years (2008 - 2017),” Human Vaccines & Immunotherapeutics, vol. 15, no. 7–8, p. 1496–1504, 2019.

[15] H. Sha, M. Al Hasan, G. Mohler and J. Brantingham, “Dynamic topic modeling of the COVID-19 Twitter narrative among U.S. governors and cabinet executives,” Association for the Advancement of Artificial Intelligence, 2020.

[16] A. Daughton, C. D Shelley, M. Barnard, D. Gerts, C. Watson Ross, I. Crooker, G. Nadiga, N. Mukundan, N. Yadira Vaquera Chavez, N. Parikh, T. Pitts and G. Fairchild, “Mining and Validating Social Media Data for COVID-19–Related Human Behaviors Between January and July 2020: Infodemiology Study,” JOURNAL OF MEDICAL INTERNET RESEARCH, 2021.

[17] S. Chen, L. Zhou, Y. Song, Q. Xu, P. Wang, K. Wang, Y. Ge and D. Janies, “A Novel Machine Learning Framework for Comparison of Viral COVID-19–Related Sina Weibo and Twitter Posts: Workflow Development and Content Analysis,” JOURNAL OF MEDICAL INTERNET RESEARCH, 2021.

[18] A. Adikari, R. Nawaratne, D. De Silva, S. Ranasinghe, O. Alahakoon and D. Alahakoon, “Emotions of COVID-19: Content Analysis of Self-Reported Information Using Artificial Intelligence,” JOURNAL OF MEDICAL INTERNET RESEARCH, 2021.

[19] N. Ahmed, S. C. Quinn, R. H. Gregory, V. S. Freimuth and A. Jamison, “Social media use and influenza vaccine uptake among White and African American adults,” Vaccine, 2018.

[20] D. Wawrzuta, M. Jaworski, J. Gotlib and M. Panczyk, “Characteristics of Antivaccine Messages on Social Media: Systematic Review,” JOURNAL OF MEDICAL INTERNET RESEARCH, vol. 23, no. 6, 2021.

[21] P. M Massey, M. D Kearney, M. K Hauer, P. Selvan, E. Koku and A. E Leader, “Dimensions of Misinformation About the HPV Vaccine on Instagram: Content and Network Analysis of Social Media Characteristics,” JOURNAL OF MEDICAL INTERNET RESEARCH, 2020.

[22] M. Skeppstedt, A. Kerren and M. Stede, “Vaccine Hesitancy in Discussion Forums: Computer-Assisted Argument Mining with Topic Models,” Building Continents of Knowledge in Oceans of Data: The Future of Co-Created eHealth, 2018.

[23] L. Zhang, H. Fan, C. Peng, G. Rao and Q. Cong, “Sentiment Analysis Methods for HPV Vaccines Related Tweets Based on Transfer Learning,” Healthcare, 2020.

[24] A. Hussain, A. Tahir, Z. Hussain, Z. Sheikh, M. Gogate, K. Dashtipour, A. Ali and A. Sheikh, “Artificial Intelligence–Enabled Analysis of Public Attitudes on Facebook and Twitter Toward COVID-19 Vaccines in the United Kingdom and the United States: Observational Study,” JOURNAL OF MEDICAL INTERNET RESEARCH, 2021.

[25] S. Wai Hang Kwok, S. Kumar Vadde and G. Wang, “Tweet Topics and Sentiments Relating to COVID-19 Vaccination Among Australian Twitter Users: Machine Learning Analysis,” JOURNAL OF MEDICAL INTERNET RESEARCH, 2021.

[26] D. M. Blei, A. Y. Ng and M. I. Jordan, “Latent Dirichlet Allocation,” Journal of Machine Learning Research, 2003.

[27] P. Bojanowski, E. Grave, A. Joulin and T. Mikolov, “Enriching Word Vectors with Subword Information,” Transactions of the Association for Computational Linguistics, 2017.

[28] M. Müller, M. Salathé and P. E. Kummervold, “Covid-twitter-bert: A natural language processing model to analyse covid-19 content on twitter,” arXiv preprint, 2020.

[29] J. M Banda, R. Tekumalla, G. Wang, J. Yu, T. Liu, Y. Ding, K. Artemova, E. Tutubalina and G. Chowell, “A large-scale COVID-19 Twitter chatter dataset for open scientific research - an international collaboration,” arXiv, 2020.

[30] J. Devlin, M.-W. Chang, K. Lee and K. Toutanova, “BERT: Pre-training of Deep Bidirectional Transformers for Language Understanding,” arXiv, 2018.

[31] L. Martin, B. Muller, P. Javier Ortiz Suárez, Y. Dupont, L. Romary, E. Villemonte de la Clergerie, D. Seddah and B. Sagot, “CamemBERT: a Tasty French Language Model,” arXiv, 2020.

[32] E. López-Medina. P. LóPEZ, I. Hurtado, D. Dávalos, O. Ramirez, E. Martínez, J. Díazgranados, J. Oñate, H. Chavarriaga, S. Herrera, B. Parra, G. Libreros, M. Torres, M. Lesmes, C. Rios and I. Caicedo, “Effect of ivermectin on time to resolution of symptoms among adults with mild COVID-19: a randomized clinical trial,” Jama, vol. 325, no. 14, pp. 1426–1435, 2021.

[33] C. Oldenburg, B. Pinsky, J. Brogdon, C. Chen, K. Ruder, L. Zhong, F. Nyatigo, C. Cook, A. Hinterwirth, E. Lebas, T. Redd, T. Porco, T. Lietman, B. Arnold and T. Doan, “ffect of oral azithromycin vs placebo on COVID-19 symptoms in outpatients with SARS-CoV-2 infection: a randomized clinical trial,” Jama, vol. 326, no. 6, pp. 490–498, 2021.

[34] P. M Massey, A. Leader, E. Yom-Tov, A. Budenz, K. Fisher and A. C. Klassen “Applying Multiple Data Collection Tools to Quantify Human Papillomavirus Vaccine Communication on Twitter,” J Med Internet Res, 2016.

[35] WHO, “Ten threats to global health in 2019,” 2019. [Online]. Available: https://www.who.int/news-room/spotlight/ten-threats-to-global-health-in-2019. [Accessed 20 June 2021].

[36] S. Taylor, C. A. Landry, M. M. Paluszek, R. Groenewoud, G. S. Rachor and G. J. G. Asmundson, “A Proactive Approach for Managing COVID-19: The Importance of Understanding the Motivational Roots of Vaccination Hesitancy for SARS-CoV2,” Front. Psychol., 2020.

[37] D. Jolley and K. M. Douglas, “The effects of anti-vaccine conspiracy theories on vaccination intentions.,” PloS one, 2014.

[38] S. Shahsavari, P. Holur, T. Wang, T. R. Tangherlini and V. Roychowdhury, “Conspiracy in the time of corona: automatic detection of emerging COVID-19 conspiracy theories in social media and the news,” Journal of Computational Social Science, vol. 3, p. 279–317, 2020.

[39] W. Audureau, “Les discours antivaccin, bien implantés en France, ont redoublé de vigueur avec la crise sanitaire,” June 2020. [Online]. Available: https://www.lemonde.fr/les-decodeurs/article/2020/06/10/pour-les-antivaccins-les-deux-themes-dominants-sont-big-brother-et-big-pharma_6042339_4355770.html. [Accessed august 2021].

[40] M. R. Mehra, S. S. Desai, F. Ruschitzka and A. N. Patel, “RETRACTED: Hydroxychloroquine or chloroquine with or without a macrolide for treatment of COVID-19: a multinational registry analysis,” The Lancet, 2020.

[41] M. R. Mehra, S. S. Desai, S. Kuy, T. D. Henry and A. N. Patel, “Retraction: Cardiovascular Disease, Drug Therapy, and Mortality in Covid-19,” N Engl J Med, pp. 2582–2582, 2020.

[42] S. Vosoughi, D. Roy and S. Aral, “The spread of true and false news online,” Science, 2018.

[43] Diseases, “The COVID-19 infodemic,” The Lancet. Infectious Diseases, vol. 20, no. 8, p. 875, 2020.

[44] T. Burki, “The online anti-vaccine movement in the age of COVID-19,” The Lancet Digital Health, vol. 2, no. 10, pp. e504–e505, 2020.

[45] D. R. Grimes, “Health disinformation & social media: The crucial role of information hygiene in mitigating conspiracy theory and infodemics.,” EMBO reports, 2020.

